# Spread of COVID-19: Investigation of universal features in real data

**DOI:** 10.1101/2020.05.20.20107797

**Authors:** Subir K. Das

## Abstract

We present results on the existence of various common patterns in the growth of the total number of patients affected by COVID-19, a disease acquired through infection by a novel coronavirus, in different countries. For this purpose we propose a scaling model that can have general applicability in the understanding of real data of epidemics. This is analogous to the finite-size scaling, a technique used in the literature of phase transition to identify universality classes. In the disease model, the size of a system is proportional to the volume of the population, within a geographical region, that have been infected at the death of the epidemic or are eventually going to be infected when an epidemic ends. Outcome of our study, for COVID-19, via application of this model, suggests that in most of the countries, after the ‘onset’ of spread, the growths are described by rapid exponential function, for significantly long periods. In addition to accurately identifying this superuniversal feature, we point out that the model is helpful in grouping countries into universality classes, based on the late time behavior, characterized by physical distancing practices, in a natural way. This feature of the model can provide direct comparative understanding of the effectiveness of lockdown-like social measures adopted in different places.

## I. INTRODUCTION

Understanding of the pattern in the spread of an epidemic [1-4] is of immense importance. This helps minimize damage via optimal imposition of lockdown-like physical distancing (PD) measures [5], before medical solutions are found. A well-known theoretical result predicts exponential behavior for the natural spread of an epidemic [3, 4, 6, 7], viz.,

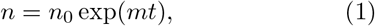

where *n* is the number of people infected till time *t*, with *n*_0_ and *m* being constants. There exist other expectations as well [4]. Power laws or even slower rates [4, 7] may not be surprising outcomes in real situations. Even if exponential, it can last only for a limited period. The late time deviation from Eq. (1) can occur due to natural reasons as well as because of imposed social restrictions.

Advanced methods of analysis [8, 9] are needed to obtain accurate picture of the overall *real* trend. There should be search for techniques that can help, for a given epidemic, identify the existence of common features, in the global scenario, for periods of ‘actual’ natural spread as well as spread during social restrictions. This is in line with the investigation of universality that is observed in phenomena associated with growth during phase transitions in materials [10-24]. Accumulation of such information has immense importance in tracking deficiencies in facilities related to medical testing as well as in identifying inadequacies in social measures. These are relevant for fighting both current and future catastrophes. Here we present results on the spread of COVID-19 [5, 7, 25–34], by applying one such model to the real data [5]. The model can have general applicability in the studies of epidemic and is related to the finite-size scaling [8, 9, 35–43], a technique used in numerical [9, 44, 45] studies to identify universality in phase transitional anomalies [10–24, 46]. The motivation behind the choice is more clearly sketched below.

Like in materials, concepts of phase transitions and universality exist in lives and societies [47, 48] as well, which are, in a sense, part of the currently popular area concerning biologically “active matter” [49–53]. Universality emerges from the fact that, if there exists some basic similarity, microscopic details do not matter [10–14, 20]. In the societal context also, as long as there exists interaction among individuals, there should exist universality in the quantitative outcomes of various phenomena, including the spread of infectious disease. This is despite differences in cultures and governments across boundaries. It is of utmost importance to quantify such feature.

A key point behind the universality in anomalous behavior is the divergence of appropriate characteristic length scale with the approach to certain fixed point [10–24]. In finite systems there is scaling of these lengths with the size of the systems, in a limiting situation [8, 9, 37, 38]. This fact is exploited in numerical studies with finite systems [8, 9, 35–38] to identify the anomalies in various quantities. Such a strategy should work for the real data on the spread of epidemic as well. In this problem, countries can be identified with materials as well as finite systems, given that the populations are nondivergent. However, unlike the standard scenario of studies in computers, where the size of a system is *a priori* assigned, here the issue is not straight-forward, particularly for an ongoing epidemic.

Outcome of our study, using real data [5], suggests that, for a large number of countries, the early time growth can be described by a prolonged “universal” exponential form, varying from country to country only via a metric factor. Various effects, including those from the practices of PD, modify this growth at late times. This is analogous to the emergence of finite-size effects [8, 37, 38]. It is shown how from the shape or form of such PD affected parts of the overall scaling function the countries can be grouped into classes in a natural way, thereby suggesting the change that may be needed in future to strengthen the PD. From academic as well as practical points of view, it is extremely encouraging and interesting to observe that the scaling concepts of statistical physics work for real data of epidemic.

## II. MODEL

In the equilibrium context, say, in critical phenomena, anomaly in a property *X*, thermodynamic or dynamic, is quantified as [10, 12-14, 20-24] *X* ~ *ϵ*^−^*^x^*, where *ϵ* is typically the deviation of the temperature of the system from the critical value and *x* is a critical exponent. The value of x is same for vastly different materials, implying universality. Note that *ξ* diverges as *ξ* ~ *ϵ*^−^*^ν^*, for thermodynamically large systems [10], so that *X* ~ *ξ^x^*^/^*^ν^*. In finite systems such divergences get restricted. This is because [8] *ξ* cannot grow beyond L, the size of the system. For *ξ* = *L*, “true” at finite-size criticality, one writes the singularity as [8, 9] *X* ~ *L^x^*^/^*^ν^*. This is the expected behavior when *X* is estimated at the “finite-size” critical points [36, 54]. The *L* = *∞* and *L* < *∞* behavior are bridged by the introduction of a scaling function *Y*(*y*) as [8, 9, 35] *X* = *Y*(*y*)*L^x^*^/^*^ν^*. Here *y* (= (*L*/*ξ*)^1/^*^ν^*) is a scaling variable that provides information on the deficiency of the size of a system with respect to the thermodynamic limit. *Y* is a constant in the *y* = 0 limit. For y → *∞*, one has *Y* ~ *y*^−^*^x^*, that is consistent with the divergence of *X* for *L* = *∞*.

In the finite-size scaling method, correct behavior of a quantity is identified by observing collapse of data [9], along with the satisfaction of the limiting behavior, from different system sizes, for *Y*. In the case of power-laws, one treats the values of the exponents as adjustable parameters in the collapse experiments.

Similar analyses [37, 38, 42, 43] have been performed for quantifying the singularities in the nonequilibrium domain. The current problem is more closely related to this. Here we briefly discuss the case of *ℓ*. This quantity diverges as *ℓ* ~ *t^α^*, where *α* is the growth exponent. In the long time limit *ℓ* = *L* and one writes for the scaling ansatz [37, 38] *ℓ* = *Y*(*y*)*L*, with *y* = (*L*/*ℓ*)^1/^*^α^* (= *L*^1/^*^α^*/*t*). In the *y* → ∞ (i.e., *L* → ∞ or *t* → 0) limit [37, 38], one should have *Y* ~ *y*^−^*^α^*. Of course, *Y* is a constant in the other limit.

As already mentioned, for the spread of epidemic, a simple theory predicts exponential growth [6]. Even for such a growth a finite-size (type) scaling equation can be constructed. Note that in the literature of coarsening also growths other than power-laws are discussed [55, 56]. For an epidemic, the size of a system should be *N*, the volume of the population that is infected when the spread stopped, i.e., the epidemic died. This number should not necessarily be proportional to the total population of a country. This statement is justifiable if the spread of COVID-19 is carefully followed [5] (see Fig. 1). It is clearly recognizable that the rate of infection is different in different countries. Thus, the final numbers may not have connection with the total population.

**FIG 1.**
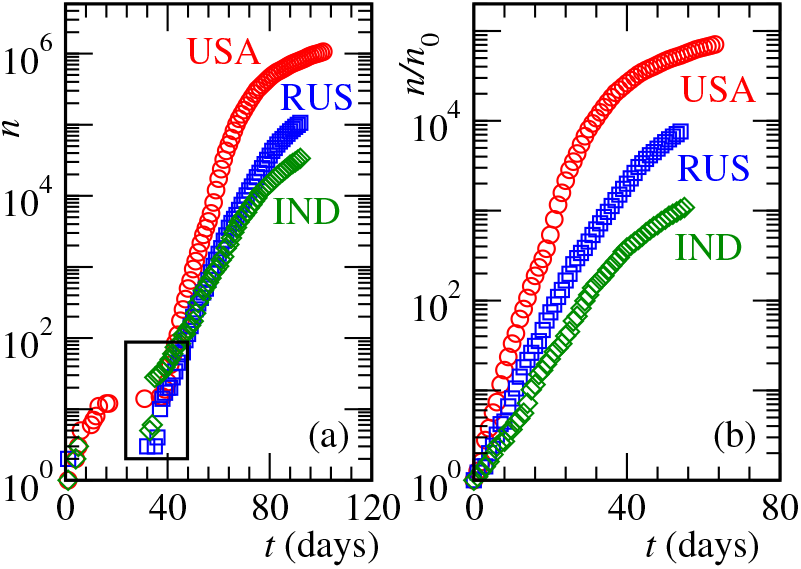
(a) Plots of *n*, the total numbers of infections, in three countries, versus time, on a semi-log scale. For each of the countries the time has been counted from the day the first infection was reported. The box at the bottom is placed to highlight the onsets of instability. (b) Similar to (a) but here the times have been counted from the days the instabilities occurred in respective countries (see text for further details). Furthermore, the ordinate, for each of the countries, has been scaled by *n*_0_, the number of infections on the day of instability.

If the rates of infection are different, one may, of course, raise question on the validity or usefulness of the approach. However, despite the differences in rate, there may still be uniqueness in the overall functional form, perhaps differing only in certain metric factors from one country to the other. The growth in Eq. (1), with differences in *m*, is one such example.

For the form in Eq. (1) one may need to adjust the constant *m*, if different for different countries, to obtain collapse of data. Here the scaling ansatz is

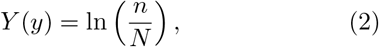

the scaling variable being

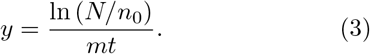

In the limit *y* → ∞, *Y* should behave as

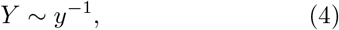

and it must approach a constant when *y* → 0.

In case an analysis is being performed prior to the death of an epidemic, the value of *N* cannot be known. It is also not expected that the finite-size behavior, i.e., the growth in the presence of PD, after the knowledge of the disease has adequately spread, will be same for all the countries. This non-unique feature will lead to lack of data collapse. However, both uniqueness and non-uniqueness in this context can provide useful information. The issues on the choice of N in the case of analysis prior to the death and non-universality in m will be discussed later. One can, of course, introduce an exponent, to check for the stretched or stressed character in the exponential function. However, we will not travel this path.

## III. RESULTS

We have worked with data for more than 15 countries. However, for the sake of brevity, we present results for a few most populous countries. These are United States of America (USA), Russia (RUS) and India (IND). The data sets contain the numbers till April 30, 2020. The first confirmed cases in these countries were reported on the following dates: USA – 21 January; RUS – January 31; and IND – January 30; all in the year 2020, of course [5].

In Fig. 1 (a) we show *n* versus t plots for the considered countries, the unit for the latter being a day. The times in these plots are counted from the dates on which the first confirmed cases were reported [5]. These plots are shown on a semi-log scale. It is clearly appreciable that n and its rate of change vary drastically from country to country [26]. Even though bending is visible, for some early period the spread may be exponential. However, confirmation of this from fitting exercise is ambiguous, particularly because of the lack of confidence in the choice of regimes, in the presence of PD effects at late times.

From the plots in Fig. 1 (a), it is also clear that in none of the countries the “instability” has set in, i.e., infections within the countries have truly started, until beyond t=30. Please see the parts put inside the box. Up to certain times, from *t* = 0, the growths that are visible occurred essentially due to arrivals of patients from abroad. It took time for the confirmations of the infections spread by these patients. This says that it is appropriate to start counting time from the day the infections from “within” a country have started getting reported. For the rest of paper, these appropriately chosen onset times (USA: 38; RUS: 38; IND: 37) have been subtracted from those used in Fig. 1 (a). In the following we have also normalized *n* by *n*_0_, the number at the onset. That way, for each country the depicted growth implies spread by starting from a single patient. This puts all the countries on fair footing at the beginning. These transformed data sets are shown on a semi-log scale in Fig. 1 (b).

The results in Fig. 1 (b) convey the message that the rate of spread of the disease in each of the countries is very different from the others. If the behavior is really exponential, the factor *m* can significantly differ among the countries. The deviations from the exponential-like behavior, at *n*/*n*_0_ = *n_d_*, after certain times, are primarily because of PD, that includes the effects of lockdown, and this fact is analogous to the appearance of the finite-size effects [15, 38]. In a standard phase transition problem, the characteristic length at the departure of a quantity from the thermodynamic limit behavior, i.e., the length at the onset of finite-size effects, is proportional to the system size [38, 42, 43]. Thus, instead of the actual size *N*/*n*_0_ of the system, here one can work with *n_d_*, value of which is country specific. Since the death of COVID-19 has not arrived yet, this is the only option we have.

In Fig. 2 we have presented results from our scaling model. Here we have shown *Y* as a function of *y*, by including data from all the considered countries. The collapse of data appears good. The corresponding coordinates of (best) parameters, (*n_d_*, *m*), for USA, RUS and IND are (2000, 0.330), (800, 0.205) and (200, 0.155), respectively. In the inset we have shown the same results on a double-log scale. Here also the collapse looks nice. The solid line in the inset is a power-law with exponent −1. A simple exponential growth will imply consistency of the data with this exponent in the large *y* limit. However, there is deviation by about 5%. While this can be due to statistical error, we do not discard the possibility of an exponential behavior with a slightly nonlinear argument.

**FIG 2.**
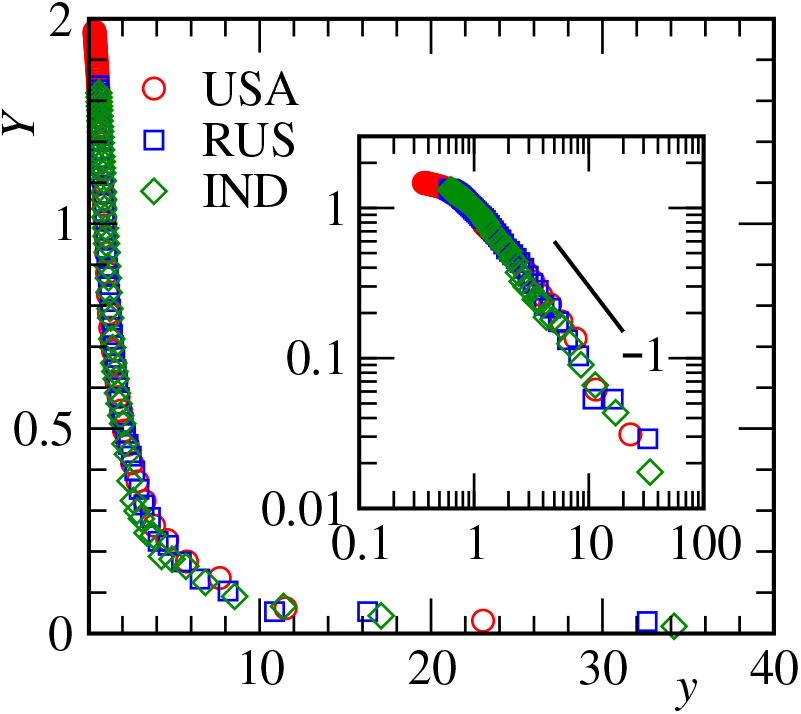
Finite-size scaling analysis, i.e., *Y* versus *y* plots, to understand the spread of COVID-19. Data from all the considered countries have been included. See text for the values of m and *n_d_* that were used to obtain the data collapse. The main frame and the inset show the same plots on linear and log scales. The solid line in the inset is a power-law. The value of the exponent has been mentioned next to it.

For the late time behavior, it is not expected that the data sets from all the countries will overlap with each other. This is because, the success of PD depends upon several factors, including economic prosperity and population density. Even if good overlap is not observed in this regime, a fair idea about relative country-wise deficiency in the effectiveness of lockdown-like social measures [5] can be obtained. This is because of the fact that the analysis has the potential of getting data from all the countries overlapped at the very least till the appearance of the effects of PD, if the natural growth in different countries are described by a unique function, apart from non-universal [39, 40] metric factors, which is ‘*m*’ for Eq. (1). Such a collapse cannot be obtained via a simple scaling by the metric factors and thus, the relative knowledge of the effectiveness of PD in different countries will remain largely unexplored.

For the presented countries, of course, we obtain very good collapse of data over the whole range. The disparities in the economic and similar parameters among these countries are well known. Nevertheless, similarity or universality in a robust way, as implied by the collapse throughout, is quite interesting, even if within a limited set of countries. In fact more countries should belong to this class. An interesting point to notice here is the following. The value of *m* for each of these countries is different from the others. A near perfect overall collapse of data, nevertheless, implies that in the post-PD regimes also these countries are consistently maintaining same discrepancy from each other as the pre-PD regimes. These are interesting facts and understanding needs attention. We have identified classes other than this. In one of those belong South Korea, Australia and few other countries. Another class is formed by most of the large West European countries.

The correctness of the numbers quoted above for *n_d_* and *m*, obtained via the optimum collapse of data from different countries, can be judged from Fig. 3 (a). There we have re-plotted the data sets of Fig. 1 (b) and compared them with the exponential form of Eq. (1) after inserting the scaling numbers for *m*. Values of *n_d_* for each of the countries can be read out from the departure points of the exponential functions from the data sets represented by symbols and compared with the above quoted numbers. The agreement is rather good. This further justifies the scaling method and the functional form for the “natural” spread of the disease. We performed the following exercise for further confirmation of the latter.

**FIG 3.**
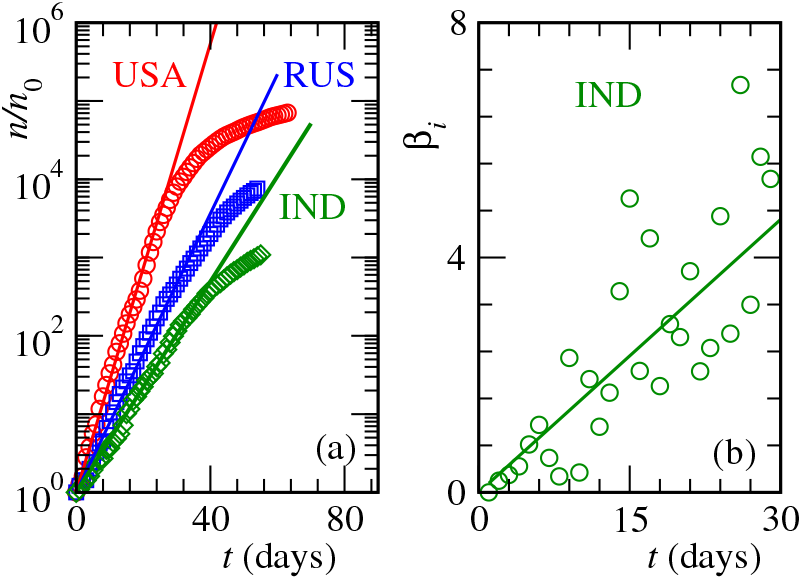
(a) Symbols represent same data sets as in Fig. 1 (b). Here we have added the exponential function of Eq. (1) (see the solid lines) by inserting the best values of *m* that were obtained from the scaling analysis. See text for these numbers. (b) The instantaneous exponent, *β_i_*, is plotted versus *t*, for IND. The solid line represents Eq. (6) with *m* equaling the scaling value for this country.

We have calculated *β_i_*, defined as [38, 42, 57–59]

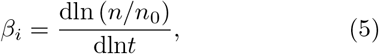

the logarithmic time derivative of the growth data in Fig. 1 (b). From Eq. (5) it is appreciable that the purpose of the quantity is to provide information on power-law [57, 58], and so, *β_i_* is referred to as the instantaneous exponent. Nevertheless, this quantity is helpful in identifying other possibilities as well [42, 59].

In Fig. 3 (b) we have presented a plot for *β_i_*, for IND, as a function of t. The PD affected region has been carefully removed. For the behavior in Eq. (1),

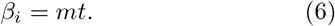

The presented data set is consistent with this linear expectation, with the scaling value of ‘*m*’ being a near perfect number for the slope. A nonlinear fit (*β*_i_ ~ *t^c^)* provides a value of the exponent close to unity, viz., *c* ≃ 1.08. The deviation from the linearity perhaps again suggests that the actual growth is slightly shifted towards a case where the argument inside the exponential is marginally nonlinear.

## IV. CONCLUSION

Finally, our study is suggestive of “prolonged” exponential growth, in all practical sense, in the natural spread of the epidemic [3, 4]. Despite differences in population density and economic parameters, it seems there is universality in the growths and the effects of physical distancing practices. Even if there exist multiple universality classes, the proposed scaling model is useful. In addition to identifying the pre-PD growth, the relative deficiencies in the measures related to PD can be well captured in the outcome. Very late time outcomes of lockdown cannot be tested by our model now. Such second order effects can be included in future.

Exponential growth is commonly related to an ideal picture of spread of rumours, where, say, every knowledgeable person spreads a hoax to one more individual every next day. But in the case of an infectious disease, various factors can resist such a spread, even before any strict social measures have been implemented. E.g., beyond a certain time either the patients get cured or they die, thereby leave the gang of spreaders. Thus, even the natural spread can be slower. If the above picture is true, from the duration of exponential growth it appears that the patients remain ill over long period. This is consistent with the medical observation. Nevertheless, we do not discard possibilities other than the above mentioned ideal picture. Continuing with the above reasoning, one should note that the patients are put under surveillance immediately after being tested positive beyond which they typically do not infect others. We expect the time gap between being infected and being tested positive to be less than the duration of exponential spread, the latter being significantly larger than 20 days in many countries. In that case, there may as well be further reasons behind this fast growth.

## Data Availability

The data are available with the author.

## Acknowledgment

The author acknowledges financial support from the Department of Biotechnology, India (grant No. LSRET-JNC/SKD/4539); and Science and Engineering Research Board of Department of Science and Technology, India (grant No. MTR/2019/001585).

